# Antibody Responses to 3^rd^ Dose mRNA Vaccines in Nursing Home and Assisted Living Residents

**DOI:** 10.1101/2021.12.17.21267996

**Authors:** Ali Zhang, Jessica A. Breznik, Rumi Clare, Ishac Nazy, Matthew S. Miller, Dawn M. E. Bowdish, Andrew P. Costa

## Abstract

A comparison of SARS-CoV-2 wild-type and the beta variant virus neutralization capacity between 2 and 3 mRNA vaccine series in nursing home residents, and between nursing home and assisted living residents strongly supports 3^rd^ dose vaccine recommendations, and equivalent polices for nursing homes and assisted living settings. Findings suggest that residents mount a robust humoral response to a 3^rd^ mRNA vaccination, and have greater neuralization capacity compared to a 2 dose series.

## Introduction

A 3^rd^ dose COVID-19 vaccination was recommended in U.S. long-term care settings on September 24, 2021^1^, though only half of eligible nursing home residents have received a 3^rd^ dose as of Dec 5 2021^2^. On August 17, 2021, the province of Ontario, Canada, was relatively early to recommend a 3^rd^ dose mRNA vaccination for residents in nursing home and assisted living settings in response to waning protection^3^. Early real-world vaccine effectiveness data suggests substantially greater protection from Omicron (B.1.1.529; hereafter Omicron) infection for persons who have received a 3^rd^ dose mRNA vaccine, compared to a 2-dose vaccination series^4^. It is unclear if residents in long-term care settings mount a robust response to a 3^rd^ mRNA vaccination that may confer protection.

## Methods

In this serial cross-sectional analysis, we examined SARS-CoV-2 neutralizing antibody titers after both 2^nd^ and 3^rd^ vaccination in nursing home residents, and after 3^rd^ vaccination in assisted living (also known as retirement home) residents from Ontario, Canada. Residents were recruited from seventeen nursing homes between March and November 2021, and eight assisted living facilities between August and November 2021. Nursing home residents were administered Moderna Spikevax 100 mcg (mRNA-1273; hereafter Moderna) or Pfizer-BioNTech Comirnaty 30 mcg (BNT163b2; hereafter Pfizer) as per recommended 2 dose schedules. Nursing home and assisted living participants were administered either Moderna or Pfizer 3^rd^ dose vaccine no earlier than 6 months post 2^nd^ dose. All protocols were approved by an ethics board, and informed consent was obtained.

Neutralization capacity of antibodies was assessed by cell culture assays with live SARS-CoV-2 virus, with data reported as geometric microneutralization titers at 50% (MNT50) which ranged from below detection (MNT50 = 10) to MNT50 = 1280^3,5^. Antibody neutralization was measured against the wild-type strain of SARS-CoV-2 and the beta variant of concern (B.1.351; hereafter beta variant). The beta variant, the most immunologically similar to omicron to date, was obtained through BEI Resources, NIAID, NIH: SARS-Related Coronavirus 2, Isolate hCoV-19/South Africa/KRISP-K005325/2020, NR-54009, contributed by Alex Sigal and Tulio de Oliveira.

Median neutralization levels by days since vaccination for the SARS-CoV-2 wild-type strain and the beta variant were plotted for comparisons. Median neutralization levels were compared by Kruskal-Wallis test for overlapping time points, and proportion below detection by chi-square, using SAS 9.4 (SAS Institute Inc.).

## Results

A total of 418 nursing home residents were recruited 12-240 days post 2^nd^ dose, with an average (standard deviation; SD) age of 82.3 (11.5) years and 63.5% (263) being female sex. The Moderna-Moderna series was administered to 52.9% (221) residents, and the Pfizer-Pfizer series to 46.4% (194) residents. A total of 103 nursing home residents were recruited 12-77 days post 3^rd^ dose, with an average (SD) age of 83.8 (10.3) years and 62.8% (64) being female sex; whereas 95 assisted living residents were also recruited 12-77days post 3^rd^ dose, with an average (SD) age of 85.0 (7.0) years and 65.3% (62) being female sex (p>0.050 for comparisons). Nursing home residents received Moderna-Moderna-Moderna (66.0%), Pfizer-Pfizer-Moderna (17.5%), and Pfizer-Pfizer-Pfizer (16.5%); whereas assisted living residents received Pfizer-Pfizer-Pfizer (61.1%), Moderna-Moderna-Moderna (22.1%), Pfizer-Pfizer-Moderna (14.7%), and Moderna-Moderna-Pfizer (2.1%).

For the equivalent 12-77 days post vaccine period, residents post 3^rd^ dose had substantially higher neuralization titers for both the wild type and beta variant (p<0.001). For beta variant neuralization, 20.8% (11) of post 2^nd^ dose residents were below the level of detection compared to none of the post 3^rd^ dose residents (p<0.001). Neutralization titers were substantially lower for beta variant neuralization (p<0.001). There were no differences in median neutralization titers between nursing home residents and assisted living residents 12-77 days post 3^rd^ dose, though 5.3% (5) of assisted living residents were below detection (p<0.05).

## Discussion

Our data strongly support 3^rd^ dose vaccine recommendations, and equivalent polices for nursing homes and assisted living settings. Consistent with non-frail populations^5^, our findings suggest that residents mount a robust humoral response to a 3^rd^ mRNA vaccination, with greater neuralization capacity compared to a 2^nd^ dose series. Continued monitoring of neutralization titers over time will determine the rate of degradation. Neutralization performance for the beta variant may not mimic performance for the omicron variant, which at time of writing remains a concern.

## Data Availability

All data produced in the present study are available upon reasonable request to the authors

## Acknowledgements

We acknowledge administrative and technical assistance from Tara Kajaks, PhD; Ahmad Rahim, MSc; Komal Aryal, MSc; Megan Hagerman; Braeden Cowbrough MSc; Lucas Bilaver; Sheneice Joseph; Angela Huynh, PhD; and Leslie Tan who were compensated for their contributions by a grant funded by the Canadian COVID-19 Immunity Task Force at McMaster University.

---

**Figure.**
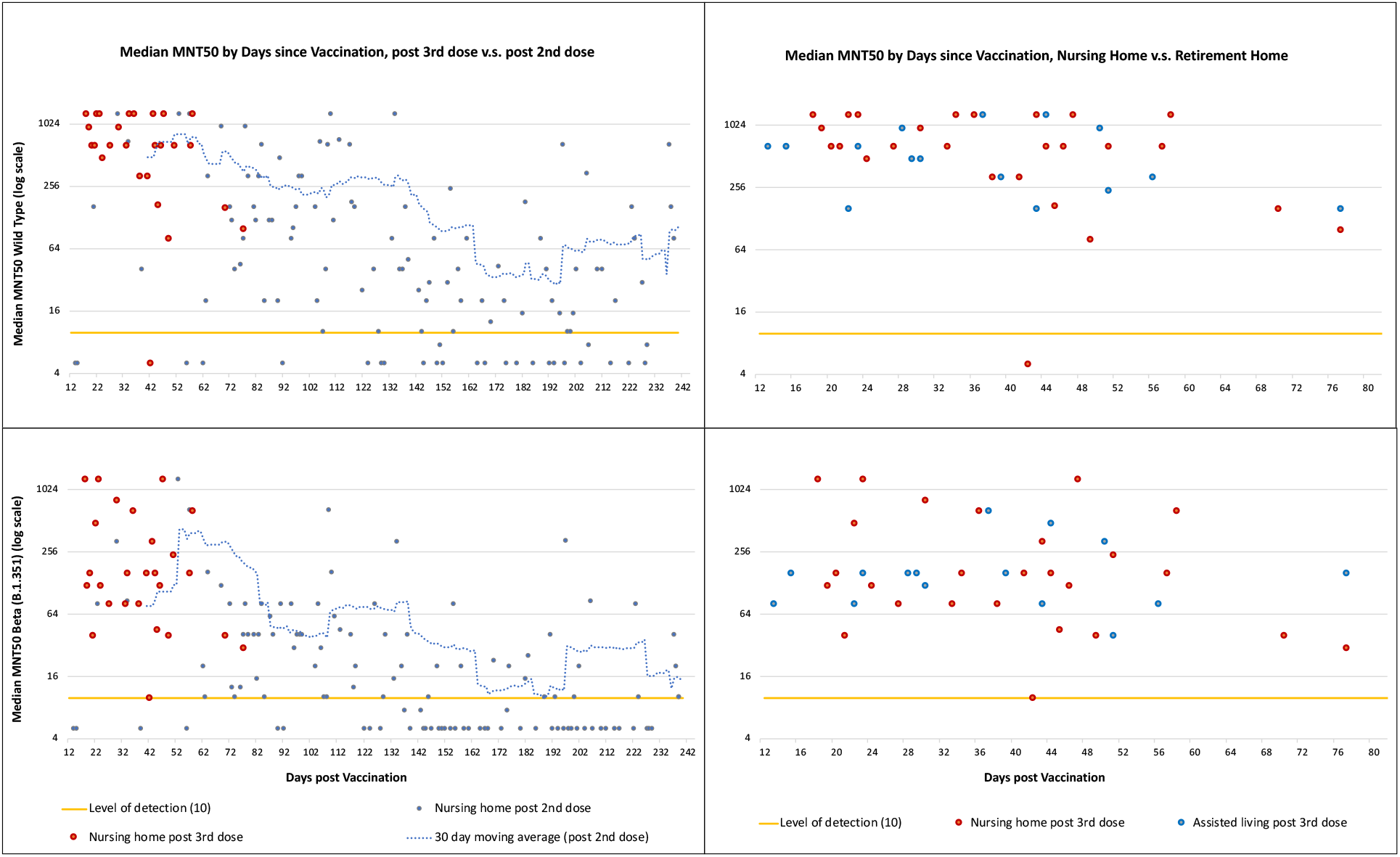
Median Neutralization (MNT50) Plots for SARS-Cov-2 Wild-Type Strain (top) and Beta Variant of Concern (B.1.351) (bottom). Nursing Home Residents post 2^nd^ compared to 3^rd^ mRNA dose (left), and post 3^rd^ mRNA dose for Nursing Home Residents compared to Assisted Living Residents (right).

