## Supplementary material for "Antibody Responses to 3^rd^ Dose mRNA Vaccines in Nursing Home and Assisted Living Residents": Methods Appendix

Study Design and Settings:

The "COVID-19 in Long-term Care Study" (https://covidinltc.ca/) is a large immunity surveillance study funded by the COVID-19 Immunity Task Force of the Public Health Agency of Canada to inform practice and immunization policy for seniors in long-term care settings. It is a prospective cohort study of nursing home residents from nursing homes and assisted living facilities (also known as retirement homes) in Ontario, Canada. Ontario is Canada's most populous province, with over 70,000 residents across 623 nursing homes and over 55,000 residents across 770 assisted living facilities. Medical and personal care in nursing homes are covered by Ontario's' universal and publicly funded health insurance plan, with residents responsible for an accommodation copayment. Assisted living facilities in Ontario are private residential complexes that provide a range of supportive care and lifestyle services that are purchased out of pocket by residents or their families.

Nursing home and assisted living residents in Ontario, Canada, were prioritized for a mass vaccination program with Moderna Spikevax 100 mcg (mRNA-1273; hereafter Moderna) or Pfizer-BioNTech Comirnaty 30 mcg (BNT163b2; hereafter Pfizer) beginning December 23, 2020 and were offered at least a first dose by February 13, 2021. Residents were recruited from seventeen nursing homes between March and November 2021, and eight assisted living facilities between August and November 2021.

Participants:

Residents eligible for recruitment were long stay, fully vaccinated with Moderna or Pfizer SARS-CoV-2 vaccines (as per manufacturer's recommended 2-dose schedules), and were (or had a substitute decisionmaker) proficient in English to understand consent. Consent to participation was sought in person by trained research coordinators employed by the participating long-term care homes, and by phone (for substitute decisionmakers) using similar communication protocols established for consent to vaccination.

Consenting nursing home residents were administered Moderna or Pfizer as per recommended 2 dose schedules between January 1$^{st}$ 2021 to November 4$^{th}$ 2021. Consenting nursing home and assisted living residents were administered either Moderna or Pfizer vaccines as a 3$^{rd}$ dose between August 24$^{th}$ 2021 and November 4$^{th}$ 2021 (and no earlier than 6 months post 2$^{nd}$ dose).

Collection and Analyses:

At enrollment, venous blood was drawn in anti-coagulant-free vacutainers for isolation of serum. 20 ml (2 Tbsp) of blood was collected in total including 2 x10 ml tubes for cell banking, 1 x 3 ml tubes for immunophenotyping and 1 x 7 ml tube for serum/soluble mediators. PBMCs and serum were cryopreserved using standard operation procedures developed by the Human Immune Testing Suite.

Measures:

Serum SARS-CoV-2 IgA/G/M against spike (S) protein and its receptor-binding domain (RBD) were measured by validated ELISA, with assay cut-off at the mean and 3 standard deviations of a pre-COVID-19 population from the same geographic region. Antibody levels and neutralization capacity were measured, and compared to a previously published convalescent cohort were used as a comparator. Data are reported as a ratio of observed optical density (OD) to the determined assay cut-off OD, with ratios above 1 considered positive.

Neutralization capacity of these antibodies was assessed by cell culture assays with live SARS-CoV-2 virus, with data reported as geometric microneutralization titers at 50% ($MNT_{50}$) which ranged from below detection ($MNT_{50}$ = 10) to $MNT_{50}$ = 1280. Antibody neutralization was measured against the wild-type strain of SARS-CoV-2 and the beta variant of concern (B.1.351; hereafter beta variant). The beta variant, the most immunologically similar variant available for omicron, was obtained through BEI Resources, NIAID, NIH: SARS-Related Coronavirus 2, Isolate hCoV-19/South Africa/KRISP-K005325/2020, NR-54009, contributed by Alex Sigal and Tulio de Oliveira.

Analyses:

Differences between antibody levels and neutralization in individuals that received mRNA-1273 or BNT163b2 were assessed by chi-square of independence (proportions), Kruskal-Wallis test (median), and student's t-test (mean). Median neutralization levels per time point for both the wild-type strain and the beta variant were plotted by days since vaccination. Post 2nd and 3rd dose plots for nursing homes residents we're overlaid for visual comparison, and a 30-day moving average was fit for post 2nd dose neutralization levels. Post 3rd dose plots for nursing homes and assisted living were also overlaid for visual comparison. The below detection ($MNT_{50}$ = 10) threshold was plotted for reference. Differences between median neutralization levels were assessed by Kruskal-Wallis test for overlapping time points, and the proportion of residents with neutralization level below detection was compared by chi-square of independence. All statistical analyses were conducted using SAS 9.4 (SAS Institute Inc.).

Ethics Approval:

All protocols were approved by the Hamilton Integrated Research Ethics Board, and informed consent was obtained (HiREB #13059).

**Appendix 1: Humoral Immunity Markers. NH post 2nd dose (12-240 days) vs NH post 3rd dose (12-77 days) vs AL post 3rd dose (12-77 days)**

| | *Population* | | | | |
|---|---|---|---|---|---|
| | *NH post 2nd dose (N = 418)* | *NH post 3rd dose (N = 103)* | *RH post 3rd dose (N = 95)* | *Total (N = 616)* | *P Value* |
| **Age** | | | | | |
| Mean (SD) | 82.30 (11.45) | 83.80 (10.28) | 84.99 (6.98) | 82.97 (10.72) | 0.06 |
| Median (IQR) | 85.0 (77.0 – 90.0) | 86.0 (78.0 – 92.0) | 85.0 (81.0 – 90.0) | 85.0 (78.0 – 91.0) | 0.37 |
| **Sex** | | | | | |
| F | 263 (063.53%) | 064 (062.75%) | 062 (065.26%) | 389 (63.67%) | 0.96 |
| **Vaccination type** | | | | | |
| Missing | 000 (000.00%) | 000 (000.00%) | 000 (000.00%) | 000 (00.00%) | |
| Moderna X2 | 221 (052.87%) | 000 (000.00%) | 000 (000.00%) | 221 (35.88%) | |
| Moderna X3 | 000 (000.00%) | 068 (066.02%) | 021 (022.11%) | 089 (14.45%) | |
| Moderna, Moderna, Pfizer | 000 (000.00%) | 000 (000.00%) | 002 (002.11%) | 002 (00.32%) | |
| Pfizer X2 | 194 (046.41%) | 000 (000.00%) | 000 (000.00%) | 194 (31.49%) | |
| Pfizer X3 | 000 (000.00%) | 017 (016.50%) | 058 (061.05%) | 075 (12.18%) | |
| Pfizer, Moderna | 003 (000.72%) | 000 (000.00%) | 000 (000.00%) | 003 (00.49%) | |
| Pfizer, Pfizer, Moderna | 000 (000.00%) | 018 (017.48%) | 014 (014.74%) | 032 (05.19%) | |
| **IgG Spike** | | | | | |
| N (N Missing) | 418 (0) | 103 (0) | 95 (0) | 616 (0) | |
| Mean (SD) | 2.11 (0.87) | 2.83 (0.30) | 2.81 (0.19) | 2.34 (0.80) | <0.001*** |
| Median (IQR) | 2.4 (1.4 – 2.9) | 2.9 (2.9 – 2.9) | 2.9 (2.8 – 2.9) | 2.8 (1.9 – 2.9) | <0.001*** |
| **=> IgG Spike cutoff (0.54871144)** | | | | | |
| Missing | 000 (000.00%) | 000 (000.00%) | 000 (000.00%) | 000 (00.00%) | 0.001** |
| Above cut-off | 390 (093.30%) | 102 (099.03%) | 095 (100.00%) | 587 (95.29%) | |
| Below cut-off | 028 (006.70%) | 001 (000.97%) | 000 (000.00%) | 029 (04.71%) | |

| | | | | | |
|---|---|---|---|---|---|
| **IgG RBD** | | | | | |
| Mean (SD) | 1.45 (1.02) | 2.40 (0.71) | 2.22 (0.82) | 1.73 (1.02) | <0.001*** |
| N (N Missing) | 418 (0) | 103 (0) | 95 (0) | 616 (0) | |
| Median (IQR) | 1.2 (0.6 – 2.4) | 2.8 (2.0 – 2.9) | 2.6 (1.6 – 2.9) | 1.8 (0.7 – 2.8) | <0.001*** |
| **=> IgG RBD cutoff (0.55056615)** | | | | | |
| Missing | 000 (000.00%) | 000 (000.00%) | 000 (000.00%) | 000 (00.00%) | <0.001*** |
| Above cut-off | 314 (075.12%) | 101 (098.06%) | 091 (095.79%) | 506 (82.14%) | |
| Below cut-off | 104 (024.88%) | 002 (001.94%) | 004 (004.21%) | 110 (17.86%) | |
| **IgM Spike** | | | | | |
| Mean (SD) | 0.34 (0.37) | 0.65 (0.71) | 0.47 (0.56) | 0.41 (0.48) | <0.001*** |
| N (N Missing) | 418 (0) | 103 (0) | 95 (0) | 616 (0) | |
| Median (IQR) | 0.2 (0.2 – 0.3) | 0.4 (0.3 – 0.7) | 0.3 (0.2 – 0.4) | 0.3 (0.2 – 0.4) | <0.001*** |
| **=> IgM Spike cutoff (0.581776245384711)** | | | | | |
| Missing | 000 (000.00%) | 000 (000.00%) | 000 (000.00%) | 000 (00.00%) | <0.001*** |
| Above cut-off | 043 (010.29%) | 029 (028.16%) | 017 (017.89%) | 089 (14.45%) | |
| Below cut-off | 375 (089.71%) | 074 (071.84%) | 078 (082.11%) | 527 (85.55%) | |
| **IgM RBD** | | | | | |
| Mean (SD) | 0.18 (0.14) | 0.23 (0.31) | 0.19 (0.23) | 0.19 (0.20) | 0.045** |
| N (N Missing) | 418 (0) | 103 (0) | 95 (0) | 616 (0) | |
| Median (IQR) | 0.1 (0.1 – 0.2) | 0.2 (0.1 – 0.2) | 0.1 (0.1 – 0.2) | 0.1 (0.1 – 0.2) | 0.06 |
| **=> IgM RBD cutoff (0.597742174381105)** | | | | | |
| Missing | 000 (000.00%) | 000 (000.00%) | 000 (000.00%) | 000 (00.00%) | 0.45 |
| Above cut-off | 008 (001.91%) | 004 (003.88%) | 002 (002.11%) | 014 (02.27%) | |
| Below cut-off | 410 (098.09%) | 099 (096.12%) | 093 (097.89%) | 602 (97.73%) | |
| **IgA Spike** | | | | | |
| Mean (SD) | 1.19 (0.89) | 2.36 (0.81) | 2.01 (0.87) | 1.51 (0.99) | <0.001*** |
| N (N Missing) | 418 (0) | 103 (0) | 95 (0) | 616 (0) | |
| Median (IQR) | 0.9 (0.5 – 1.7) | 2.9 (1.7 – 2.9) | 2.2 (1.2 – 2.9) | 1.3 (0.6 – 2.7) | <0.001*** |
| **=> IgA Spike cutoff (0.543751689499111)** | | | | | |

| | | | | | |
|---|---|---|---|---|---|
| Missing | 000 (000.00%) | 000 (000.00%) | 000 (000.00%) | 000 (00.00%) | <0.001*** |
| Above cut-off | 297 (071.05%) | 100 (097.09%) | 088 (092.63%) | 485 (78.73%) | |
| Below cut-off | 121 (028.95%) | 003 (002.91%) | 007 (007.37%) | 131 (21.27%) | |
| **IgA RBD** | | | | | |
| Mean (SD) | 0.50 (0.55) | 0.83 (0.83) | 0.64 (0.73) | 0.58 (0.64) | <0.001*** |
| N (N Missing) | 418 (0) | 103 (0) | 95 (0) | 616 (0) | |
| Median (IQR) | 0.3 (0.2 – 0.5) | 0.5 (0.3 – 1.0) | 0.3 (0.2 – 0.7) | 0.3 (0.2 – 0.6) | <0.001*** |
| **=> IgA RBD cutoff (0.56369917862878)** | | | | | |
| Missing | 000 (000.00%) | 000 (000.00%) | 000 (000.00%) | 000 (00.00%) | <0.001*** |
| Above cut-off | 097 (023.21%) | 046 (044.66%) | 027 (028.42%) | 170 (27.60%) | |
| Below cut-off | 321 (076.79%) | 057 (055.34%) | 068 (071.58%) | 446 (72.40%) | |
| **Nucleocapsid IgG** | | | | | |
| N (N Missing) | 418 (0) | 103 (0) | 95 (0) | 616 (0) | |
| Mean (SD) | 0.34 (0.47) | 0.31 (0.31) | 0.18 (0.15) | 0.31 (0.41) | 0.003** |
| Median (IQR) | 0.2 (0.1 – 0.3) | 0.2 (0.1 – 0.4) | 0.1 (0.1 – 0.2) | 0.2 (0.1 – 0.3) | <0.001*** |
| **IgG Nucleocapsid cut-off (0.5477799)** | | | | | |
| Missing | 000 (000.00%) | 000 (000.00%) | 000 (000.00%) | 000 (00.00%) | 0.014** |
| Above cut-off | 064 (015.31%) | 016 (015.53%) | 004 (004.21%) | 084 (13.64%) | |
| Below cut-off | 354 (084.69%) | 087 (084.47%) | 091 (095.79%) | 532 (86.36%) | |
| **Nucleocapsid IgA** | | | | | |
| N (N Missing) | 418 (0) | 103 (0) | 95 (0) | 616 (0) | |
| Mean (SD) | 0.23 (0.30) | 0.26 (0.31) | 0.14 (0.10) | 0.22 (0.28) | 0.007** |
| Median (IQR) | 0.2 (0.1 – 0.2) | 0.2 (0.1 – 0.3) | 0.1 (0.1 – 0.2) | 0.2 (0.1 – 0.2) | <0.001*** |
| **IgA Nucleocapsid cut-off (0.5779821)** | | | | | |
| Missing | 000 (000.00%) | 000 (000.00%) | 000 (000.00%) | 000 (00.00%) | 0.043** |
| Above cut-off | 019 (004.55%) | 006 (005.83%) | 000 (000.00%) | 025 (04.06%) | |
| Below cut-off | 399 (095.45%) | 097 (094.17%) | 095 (100.00%) | 591 (95.94%) | |
| **MNT50 (Wild type)** | | | | | |
| N (N Missing) | 418 (0) | 103 (0) | 95 (0) | 616 (0) | |

| | | | | | |
|---|---|---|---|---|---|
| Mean (SD) | 222.85 (371.45) | 719.27 (480.73) | 691.74 (493.04) | 378.17 (469.16) | <0.001*** |
| Median (IQR) | 40.0 (10.0 – 160.0) | 640.0 (320.0 – 1280.0) | 640.0 (320.0 – 1280.0) | 160.0 (20.0 – 640.0) | <0.001*** |
| **Neutralization (MNT50 Wild type)** | | | | | |
| Missing | 000 (000.00%) | 000 (000.00%) | 000 (000.00%) | 000 (00.00%) | <0.001*** |
| Above level (=>10) of detection (Wild type) | 330 (078.95%) | 102 (099.03%) | 094 (098.95%) | 526 (85.39%) | |
| Below level (<10) of detection (Wild type) | 088 (021.05%) | 001 (000.97%) | 001 (001.05%) | 090 (14.61%) | |
| **MNT50 (beta type)** | | | | | |
| N (N Missing) | 418 (0) | 103 (0) | 95 (0) | 616 (0) | |
| Mean (SD) | 58.89 (154.56) | 328.93 (396.48) | 277.53 (356.37) | 137.76 (273.95) | <0.001*** |
| Median (IQR) | 20.0 (5.0 – 40.0) | 160.0 (80.0 – 320.0) | 160.0 (40.0 – 320.0) | 40.0 (5.0 – 160.0) | <0.001*** |
| **Neutralization (MNT50 Beta type)** | | | | | |
| Missing | 000 (000.00%) | 000 (000.00%) | 000 (000.00%) | 000 (00.00%) | <0.001*** |
| Above level (=>10) of detection (Beta type) | 259 (061.96%) | 103 (100.00%) | 090 (094.74%) | 452 (73.38%) | |
| Below level (<10) of detection (Beta type) | 159 (038.04%) | 000 (000.00%) | 005 (005.26%) | 164 (26.62%) | |

***Note:***

** P value less than 0.05*

*** P value less than 0.01*

**** P value less than 0.001*

**Appendix 2: NH 2nd dose (12-77 days) vs NH 3rd dose (12-77 days)**

| | *Population* | | | |
|---|---|---|---|---|
| | *NH post 2nd dose (N = 53)* | *NH post 3rd dose (N = 103)* | *Total (N = 156)* | *P Value* |
| **MNT50 (Wild type)** | | | | |
| **N (N Missing)** | **53 (0)** | **103 (0)** | **156 (0)** | |
| **Median (IQR)** | **160.0 (40.0 – 640.0)** | **640.0 (320.0 – 1280.0)** | **640.0 (160.0 – 1280.0)** | **<0.001***** |
| **Neutralization (MNT50 Wild type)** | | | | |
| **Missing** | **000 (000.00%)** | **000 (000.00%)** | **000 (00.00%)** | **0.006**** |
| **Above level (=>10) of detection (Wild type)** | **047 (088.68%)** | **102 (099.03%)** | **149 (95.51%)** | |
| **Below level (<10) of detection (Wild type)** | **006 (011.32%)** | **001 (000.97%)** | **007 (04.49%)** | |
| **MNT50 (beta type)** | | | | |
| **N (N Missing)** | **53 (0)** | **103 (0)** | **156 (0)** | |
| **Median (IQR)** | **40.0 (10.0 – 80.0)** | **160.0 (80.0 – 320.0)** | **80.0 (40.0 – 320.0)** | **<0.001***** |
| **Neutralization (MNT50 Beta type)** | | | | |
| **Missing** | **000 (000.00%)** | **000 (000.00%)** | **000 (00.00%)** | **<0.001***** |
| **Above level (=>10) of detection (Beta type)** | **042 (079.25%)** | **103 (100.00%)** | **145 (92.95%)** | |
| **Below level (<10) of detection (Beta type)** | **011 (020.75%)** | **000 (000.00%)** | **011 (07.05%)** | |

**Note:**
*** P value less than 0.05**
**** P value less than 0.01**
***** P value less than 0.001**

**Appendix 3: AL 3rd dose (12-77 days) vs NH 3rd dose (12-77 days)**

| | *Population* | | | |
|---|---|---|---|---|
| | *NH post 3rd dose (N = 103)* | *AL post 3rd dose (N = 95)* | *Total (N = 198)* | *P Value* |
| **MNT50 (Wild type)** | | | | |
| N (N Missing) | 103 (0) | 95 (0) | 198 (0) | |
| Median (IQR) | 640.0 (320.0 – 1280.0) | 640.0 (320.0 – 1280.0) | 640.0 (320.0 – 1280.0) | 0.69 |
| **Neutralization (MNT50 Wild type)** | | | | |
| Missing | 000 (000.00%) | 000 (000.00%) | 000 (00.00%) | 1 |
| Above level (=>10) of detection (Wild type) | 102 (099.03%) | 094 (098.95%) | 196 (98.99%) | |
| Below level (<10) of detection (Wild type) | 001 (000.97%) | 001 (001.05%) | 002 (01.01%) | |
| **MNT50 (beta type)** | | | | |
| N (N Missing) | 103 (0) | 95 (0) | 198 (0) | |
| Median (IQR) | 160.0 (80.0 – 320.0) | 160.0 (40.0 – 320.0) | 160.0 (40.0 – 320.0) | 0.36 |
| **Neutralization (MNT50 Beta type)** | | | | |
| Missing | 000 (000.00%) | 000 (000.00%) | 000 (00.00%) | 0.024** |
| Above level (=>10) of detection (Beta type) | 103 (100.00%) | 090 (094.74%) | 193 (97.47%) | |
| Below level (<10) of detection (Beta type) | 000 (000.00%) | 005 (005.26%) | 005 (02.53%) | |

Note:
* P value less than 0.05
** P value less than 0.01
*** P value less than 0.001
